# Machine Learning Assisted Intraoperative Assessment of Brain Tumor Margins Using HRMAS NMR Spectroscopy

**DOI:** 10.1101/2020.02.24.20026955

**Authors:** Doruk Cakmakci, Emin Onur Karakaslar, Elisa Ruhland, Marie-Pierre Chenard, Francois Proust, Martial Piotto, Izzie Jacques Namer, A. Ercument Cicek

## Abstract

Complete resection of the tumor is important for survival in glioma patients. Even if the gross total resection was achieved, left-over micro-scale tissue in the excision cavity risks recurrence. High Resolution Magic Angle Spinning Nuclear Magnetic Resonance (HRMAS NMR) technique can distinguish healthy and malign tissue efficiently using peak intensities of biomarker metabolites. The method is fast, sensitive and can work with small and unprocessed samples, which makes it a good fit for real-time analysis during surgery. However, only a targeted analysis for the existence of known tumor biomarkers can be made and this requires a technician with chemistry background, and a pathologist with knowledge on tumor metabolism to be present during surgery. Here, we show that we can accurately perform this analysis in real-time and can analyze the full spectrum in an untargeted fashion using machine learning. We work on a new and large HRMAS NMR dataset of glioma and control samples (n = 568), which are also labeled with a quantitative pathology analysis. Our results show that a random forest based approach can distinguish samples with tumor cells and controls accurately and effectively with a mean AUC of 85.6% and AUPR of 93.4%. We also show that we can further distinguish benign and malignant samples with a mean AUC of 87.1% and AUPR of 96.1%. We analyze the feature (peak) importance for classification to interpret the results of the classifier. We validate that known malignancy biomarkers such as creatine and 2-hydroxyglutarate play an important role in distinguish tumor and normal cells and suggest new biomarker regions. The code is released at http://github.com/ciceklab/HRMAS_NC.

## 1 Introduction

Gliomas constitute 60% of all primary brain tumors [26]. The maximum resection of the tumor remains the key point in the management of gliomas with a direct influence on the survival of patients [23]. The progress made over the last two decades in surgical techniques including microsurgery by the operating microscope, preoperative functional imaging (e.g., functional MRI, MRI tractography), intraoperative electrical stimulation in awakened craniotomy and intraoperative imaging (surgery guided by real-time imaging using neuronavigation or intraoperative MRI) have largely contributed to significantly increase resected tumor volume while improving morbidity and mortality [33].

Providing feedback on left-over malign tissue during surgery can help surgeons delineate more precisely the limits of a tumor infiltration, especially after a macroscopically complete excision. Several innovative techniques based on optical spectrometry [5,6,11,14,15,17,20,24,25,29,34,36,41] or mass spectrometry [2–4, 7, 8, 13, 28, 31, 32] are now proposed to help surgeons to evaluate the margins of resection and possibly to amplify the surgical procedure. Metabolic profiling of a biopsy sample by High Resolution Magic Angle Spinning Nuclear Magnetic Resonance (HRMAS NMR) spectroscopy is a recent novel technique for efficiently distinguishing malign and healthy tissues in excision cavity during surgery. This technique is particularly well-suited for this task due to its ability to analyze small samples of unprocessed tissue specimens. It has a nondestructive nature and allows other analytical techniques on the same specimen which is important when small amounts of tissue are available [9]. Moreover, the preparation of biopsy samples is fast as it does not require lengthy chemical extraction procedures. Battini *et al*. showed that HRMAS NMR spectroscopy using intact tissue provides solid information in the characterization of pancreatic adenocarcinoma and also on the long-term survival. The information can be obtained in twenty minutes during surgery [1]. A recently released metabolic database on HRMAS NMR signatures of seventy six biomarker metabolites has taken the next step in widening the usage of the technique [30].

One challenge to overcome for this technique to be used in the surgery room is its dependence on human experts with background on chemistry and cancer biology. The raw NMR signal is evaluated by the NMR technician who can report on the existence of certain biomarker metabolites usually with no insight on the tumor metabolism. Evaluation of the raw signal comes with several obstacles. First, the identification of biomarker metabolites might not be possible due to superimposed signature signals of certain metabolites (e.g., creatine and lysine [16]). Second, certain peaks might shift due to experimental conditions (e.g., due to temperature) and then an informed guess on whether that peak belongs to the targeted metabolite must be made. Third, the intra-tumor heterogeneity might result in a convoluted signal and might make it hard for the technician to detect malignant tissue due to unusual relative peak intensities. Moreover, an expert pathologist needs to be present at the time of the surgery to relate the findings of the technicians to the tumor metabolism. Maybe the most restricting factor of this analysis pipeline is the targeted analysis of the raw NMR signal. This means the human expert is limited by the knowledge of certain biomarker metabolites and their corresponding peaks. However, the spectrum contains many uncharacterized regions which might harbor peaks that are capable of distinguishing tumor cells and yet are unknown.

In this study, we propose using machine learning approaches to address the above-mentioned problems and to automate distinguishing healthy tissue from benign/malignant tumor tissue obtained from the excision cavity during tumor surgery. The algorithm is fast and can work within the time frame of surgery. It directly outputs whether a sample includes tumor cells. Thus, it does not require a technician to analyze the signal. It performs an untargeted analysis of the signal and is able to extract information from uncharacterized regions in the spectrum. The system figure representing the proposed pipeline is shown in Figure 1.

**Fig. 1:**
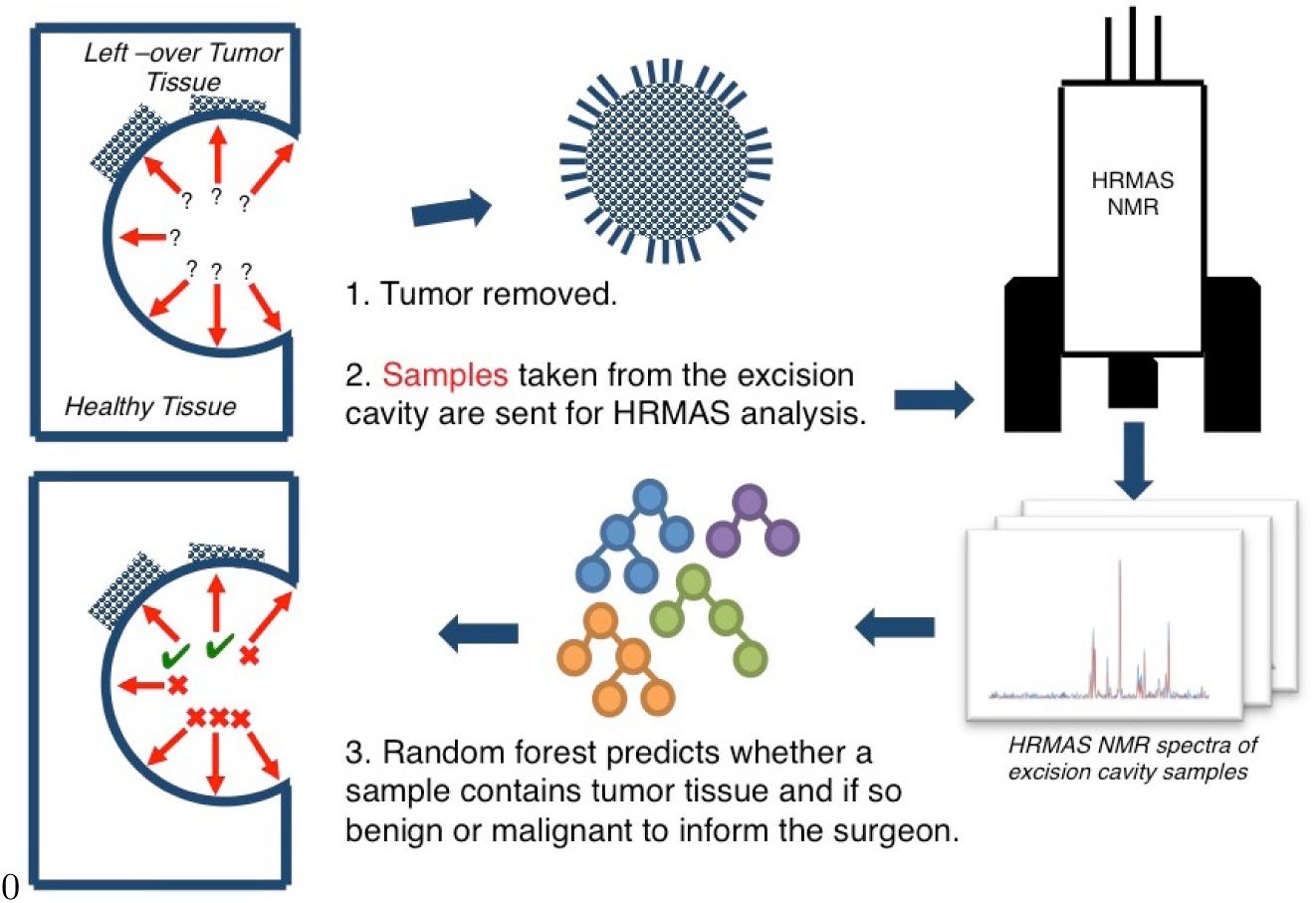
The figure shows the pipeline proposed for machine learning assisted tumor margin assessment during brain tumor surgery. After the tumor removal, the surgeon resects samples from the excision cavity. Samples are analyzed via HRMAS NMR technique. Produced spectra are processed via a random forest classifier to label each region in the cavity (malignant/benign tumor vs healthy tissue). The feedback is sent to the surgeon for resecting more tissue for regions labeled positive for tumor tissue.

Here, we utilize a new dataset (*n*=568) of glioma and control samples analyzed using HRMAS NMR. All samples are also analyzed by a clinical pathologist and labeled whether it is normal, benign or malignant. To the best of our knowledge, this is the largest of its kind with pathological labels. We benchmark various machine learning architectures and show that it is possible to distinguish tumor and control samples with a mean AUC of 85.2% and AUPR of 93.4%. We show that we can also distinguish benign and malignant tumor samples with a mean AUC of 87.1% and AUPR of 96.1%. The best performing method is a random forest based approach. This method for the first time performs an untargeted analysis of the spectrum. Moreover, the model is interpretable and informs the user about the ranges in the spectrum that were most informative for the classification, using SHAP values of the features which quantify their importance [22]. We validate that the model focuses on known cancer biomarker metabolites such as creatine and 2-hydroxylglutarate while distinguishing benign and malignant glioma samples. We also observe that branched chain amino acids have been important in the classification. We find evidence in literature that indeed altered branch chain amino-acid concentrations are related to glioma metabolism, yet, their statuses as biomarkers are not well-established. We also find some uncharacterized regions in the spectrum that are informative, which brings up further research questions on establishing an understanding on the compounds in those regions and their relation to tumor metabolism.

## 2 Materials and Methods

### 2.1 Dataset

In this subsection, we provide details on the glioma HRMAS NMR dataset and corresponding quantitative pathological analysis to obtain the labels.

#### 2.1.1 Patient’s Cohort and Tissue Sample Collection

The metabolomics-based statistical model was constructed from spectra of 400 primary brain tumor samples from 382 patients and 87 non-tumor brain tissue samples from epilepsy surgery of 73 patients. The histopathological classification of primary brain tumors is: Pilocytic astrocytoma (n=3), astrocytoma grade II (n=6), astrocytoma grade III (n=5), glioblastoma (n=189), oligodendroglioma grade II (n=40), oligodendroglioma grade II-III (n=17), oligodendroglioma grade III (n=90), oligoastrocytoma grade II (n=3), oligoastrocytoma grade II-III (n=1), oligoastrocytoma grade III (n=9), ganglioglioma grade II (5), ganglioglioma grade III (4), dysembryoplastic neuroepithelial tumors (DNET, n=26). The study on tumor margins was then performed on 271 operative samples obtained from the excisional banks of 89 patients.

Tissue specimens were collected with minimum ischemic delays after resection (average time 2 min *±*1 min), either by a pneumatic system connected between the operating theater of neurosurgery and the NMR room (Hautepierre Hospital - University Hospitals of Strasbourg), or by samples stored in two Tumor Bio-bank, Strasbourg and Colmar (Ethics Committee no. 2003-100, 09.12.2003 and no. 2013-37, 12.11.2013). A written informed consent was obtained from all patients included.

All tissue samples used in this study had a viable tumor/necrosis ratio and were quantitatively and qualitatively adequate to perform satisfactory NMR HRMAS analysis. In order to wait for this goal, after NMR HRMAS analysis, the inserts were cut, and for half the content of each sample, the percentage of tumor cells in the total sample of cells with regard to the total surface were calculated based on frozen hematoxylin & eosin-stained sections. See Supplementary Table 1 for details on collected samples.

#### 2.1.2 HRMAS NMR Data Acquisition

Each brain biopsy sample was prepared at −20°C by introducing a 15- to 18-mg biopsy into a disposable 30*µL* KelF insert. To provide a lock frequency for the NMR spectrometer, 10*µL* of *D*_2_*O* was also added to the insert.

All HRMAS NMR spectra were acquired on a Bruker (Karlsruhe, Germany) Avance III 500 spectrometer operating at a proton frequency of 500.13 MHz and equipped with a 4-mm triple-resonance gradient HRMAS probe (1H, 13C and 31P). The temperature was maintained at 4°C throughout the acquisition time in order to reduce the effects of tissue degradation during the spectrum acquisition. A one-dimensional (1D) proton spectrum using a Carr-Purcell-Meiboom-Gill (CPMG) pulse sequence was acquired with a 285*µs* inter-pulse delay and a 10-min acquisition time for each tissue sample. The number of loops was set at 328, giving the CPMG pulse train a total length of 93 ms.

### 2.1.3 HRMAS NMR Data Preprocessing

The free induction decay (FID) signal for each sample had a length of 16,384. The signal is left-shifted by 70 points to remove the Bruker digital filter in the prefix. Obtained raw FID spectrum is then transformed to frequency domain and is phased corrected. The suffix of the signal which contained almost no variance is cropped to obtain the final signal used for analysis, which is of length 8,172. The magnitude of the signal is used for the presented analysis.

### 2.2 Problem Formulation

In this study, our main task is distinguishing tumor tissue from the normal tissue. The problem is modelled as a binary classification task. For a given HRMAS NMR signal *i* in the sample set *S*, the feature vector *x*_*i*_ is a *d*-dimensional vector: 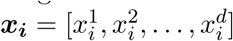 which represents the signal intensity at each ppm. The label for that sample is *y*_*i*_ and *y*_*i*_ = 1 if sample contains tumor tissue and *y*_*i*_ = 0, otherwise. Then, the model we learn is a function *f* such that *f* (***x***_***i***_) = *ŷ*_*i*_, ∀_*i*_ ∈ *S*. The second and optional task is to distinguish benign and malignant tumor samples. In this task, a sample *j* has label *z*_*j*_ = 1, if the sample has malignant tumor cells, and *z*_*j*_ = 0, if sample contains benign tumor cells. This task is also a binary classification task and we learn a function *g* such that 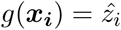. We would like to note that we also considered a multi-class classification task which unites the above mentioned binary classification tasks. However, as we discuss in Section 4, we obtained better performance with two separate tasks. Given the first task is of utmost importance and the second is optional, we opted for this approach.

### 2.3 Learning Algorithms

In this section, we describe the methods employed for the problems formulated in Section 2.2. We benchmark various machine learning algorithms to find the one suitable for the tasks at hand given the size and nature of the 1H HRMAS NMR signal. For all methods, the only input is ***x***_***i***_ for both tasks (*d* = 8, 172). See Section 3.1 for parameter details of each approach.

First, we run partial least squares discriminant analysis (PLS-DA) as a baseline which is a common method used in metabolomics analysis [40]. As the second algorithm, we used a Random Forest (RF) classifier, which trains many weak classifier trees on sample subsets which are created using bootstrapping [18] and results are aggregating via majority voting. The third algorithm is a support vector machine classifier (SVM). We employed linear and radial basis function (RBF) kernels with a soft margin.

As a baseline neural network architecture, we employ a fully connected multi-layer perceptron. Our MLP [27] model takes ***x***_***i***_ and applies a number of fully-connected (FC) layers which makes use of rectified linear unit (ReLU) activation. We use full batch gradient descent for training. At the output layer, we use softmax to assign probabilities to each class (e.g., benign) and focal loss as our loss function to address the class imbalance in our dataset (i.e., smaller number of benign samples) [19].

Convolutional neural networks (CNN) are well-established architectures for learning complex patterns on 2D image data. CNNs have also proven useful for processing 1D data. Some examples are drug chemical structure representation (e.g., SMILES [10, 37]), natural language (i.e., sentences [38]) and EEG signals [35]. Thus, we conjectured that a CNN is a good candidate for the classification tasks mentioned above. As our final model, we use a 1D CNN-based architecture to process ***x***_***i***_. The architecture consists of 2 layers of convolutional operations. First, *C*_1_ kernels of size *k*x1 are passed over the signal with stride *s* and dilation rate *d* (no padding). The same set of operations are applied on the output of the first convolutional layer with *C*_2_ kernels. The output is passed through a set of fully connected layers to produce class probabilities using softmax at the output layer. Again, we use full-batch gradient descent as our optimizer and focal loss as our loss function.

## 3 Results

### 3.1 Experimental Setup

We label the samples in our dataset (see Section 2.1) as aggressive, benign or control using the following method. Per all individuals in the dataset, we have multiple types of samples that originate from (i) the glioma tumor tissue (i.e., glioma), (ii) the healthy brain tissue (i.e., control), and (iii) from the excision cavity (i.e., test). For samples in (i), the aggressive label and the benign label are assigned with respect to the pathological analysis result. For samples in (ii), control label is assigned. For samples in (iii), if the pathology report indicates that tumor cells exist (i.e., positive), then aggressive label is assigned if the tumor of that individual is aggressive and benign label is assigned if the tumor of that individual is benign; otherwise, control label is assigned. In the end, we obtained 179 control, 88 benign and 301 malignant samples. See Supplementary Table 1 for details on the labels for collected samples. We generate 2 datasets for the two tasks explained in Section 2.2. The first one unites the labels benign and malignant and sets their labels to tumor for task 1. The second one only retains the benign and malignant samples for task 2.

Performance of the proposed models are assessed using a stratified and grouped 8-fold cross validation approach on each dataset. Each dataset is shuffled before the folds are generated. Folds are generated in a stratified manner by sampling from the dataset according to the label distribution of the dataset. That is, each fold has a similar distribution of labels to the whole dataset. There is no sample or patient overlap between the generated folds. That is, an individual’s all samples are always in a single fold and the folds are exclusive. In each iteration, first, the test and validation folds are removed. The models are trained on 6 remaining folds and the best performing parameter set is found on the validation fold. Then, each model is trained on 7 folds (training + validation) and is tested on the test fold. This procedure is repeated three times for each task with a random weight initialization of the models. AUC, AUPR distributions are calculated using the performance for each test on each test fold.

For the PLS-DA approach we used 30 components which sets the number of latent variables. For the SVM model we performed a grid search on the soft-margin regularization parameter (i.e., *C*: 0.01, 0.1, 1, 10, 100) and on the kernel choice (i.e., RBF vs linear). For the RF model, we performed a search on (i) number of estimators: 100, 300, 500, 800, and 1200; (ii) maximum tree depth: 5, 10, 15, 20, 25, and 30; and finally, (iii) minimum number of samples to split a node: 2, 5 10, 15, and 20. We also set the minimum number of samples in a leaf node to 10 to avoid overfitting. For the 4-layered fully-connected (baseline) network, the input layer has 8,127 neurons, the second layer has 4,000 neurons, the third layer has 1,000 neurons and the output layer has 2 neurons, which uses softmax to produce probabilities per class in both tasks. ReLU activation is used for all hidden layers. Finally, for the CNN model, we use two convolutional layers such that the number of kernels in both layers are *C*_1_ = *C*_2_ = 4. These 1D kernels are of size 16, 32, 64, and 128. We set stride and dilation to 1. After passing through maxpool operations of size 1×4, and ReLU activation, concatenated activation maps are input to fully-connected layers which are of size 8,112, 4,000 and 1,000, respectively. Similar to the base neural network model, output layer has 2 nodes with a softmax operation to produce class probabilities and ReLU activation is used for all hidden layers. We trained the networks with a fixed epoch number of 200, which was decided on the validation folds.

### 3.2 Performance Comparison and the Model of Choice

We compare the performances of the above-mentioned methods using AUC and AUPR metrics. Please see Figure 2a for results. For the first task, distinguishing the tumor (glioma) and control cells, all methods perform well and the lowest mean AUC achieved is 78.9% and the lowest mean AUPR achieved is 87.7%. We observe that the RF model has the best mean AUC value with 85.6% which is ∼ 1% improvement over the closest performance by the CNN model. The AUC variance of the RF model is similar to CNN and PLS-DA and smaller than other models. Similarly RF is the best performing model with respect to the AUPR metric with an AUPR of 93.4%. The second best mean AUPR is 92.6% and is achieved by CNN model. CNN model has the lowest AUPR variance and RF is the second best. In conclusion, CNN also performs almost as well as RF for this task and is slightly edged by the RF model. RF is a less complex model than CNN and more interpretable compared to CNN. Thus, it is our method of choice for this task.

**Fig. 2:**
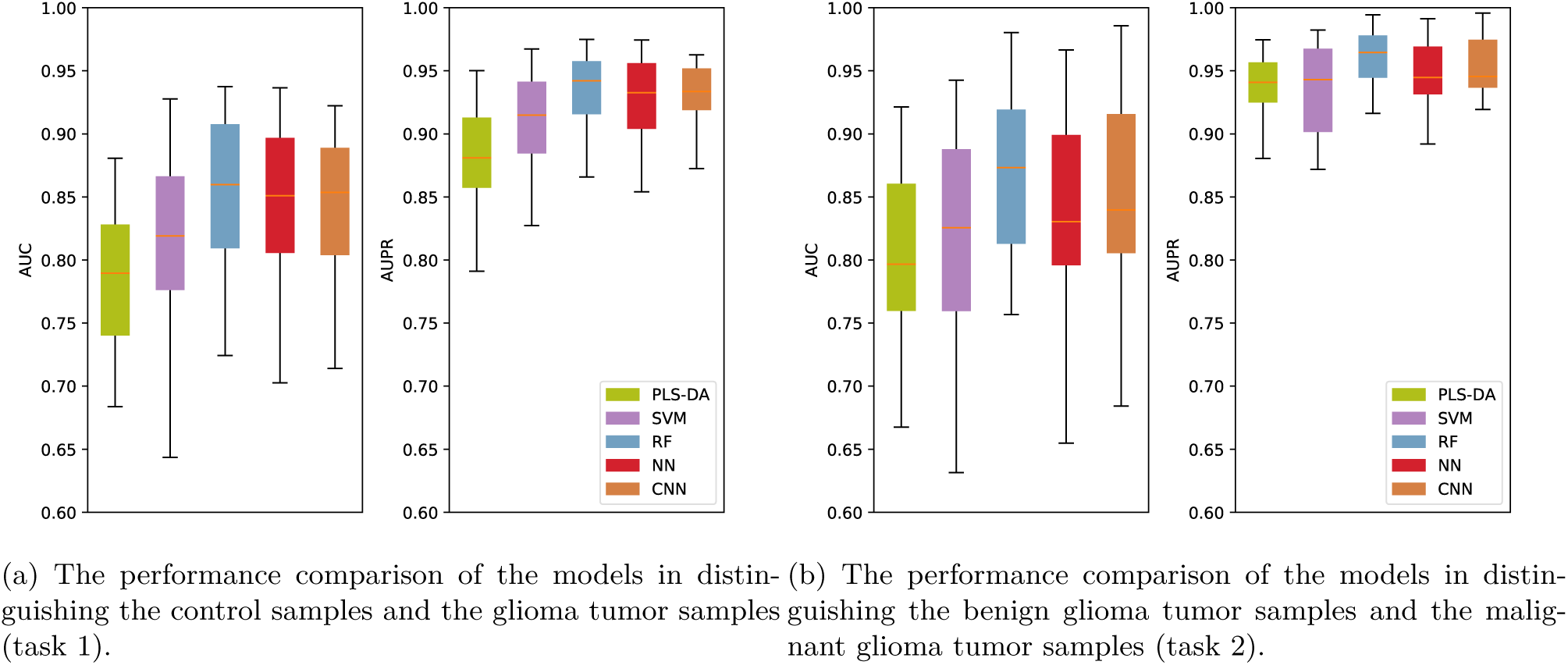
The performance comparison of the benchmarked machine learning models with respect to the AUC and AUPR metrics. Box plots represent the performance of the models obtained on the test folds, in an 8-fold cross validation setting which is repeated 3 times.

The second task in our pipeline is optional and is performed when the surgeon also would like to know if the tumor is benign or malignant. Results are shown in Figure 2b. Again, all methods perform well and the lowest mean AUC achieved is 80% and the lowest AUPR achieved is 93.4%. We observe that the RF model has the best mean AUC value with 87.1% which is ∼ 2% improvement over the closest performance by the CNN model. We also see that the RF model has the lowest AUC variance among all models. Similarly RF is the best performing model with respect to the AUPR metric with an AUPR of 96.1%. All other methods have a mean AUPR of 94%, thus also in this category RF provides a ∼ 2% improvement. The AUPR variance of RF is the lowest and is on par with CNN. Thus, RF is the model of choice for this task as well because of its robustness and high sensitivity and specificity.

### 3.3 Interpreting the Model Predictions

We analyze the feature (i.e., ppm) importance of the features that lead to correct classification of the samples in each task with the RF model. For this purpose, we make use of the SHapley Additive exPlanation (SHAP) values for each feature [21,22]. This approach has its roots in the Shapley values from coalitional game theory. Here, the features are players in a coalition and their values indicate a fair weight that represent their contribution (i.e., success of the classification.)

Here, after running the RF model for both tasks, we compute the SHAP values of each feature (i.e., ppm in the signal) for each task. Here, we map all features back to the the ppm spectrum (x-axis) and show the corresponding SHAP values (y-axis) for each sample. Each dot on this figure denotes a sample and the color of the sample denotes the value of the corresponding feature. That is, if a sample is purple it means its feature value is high, and if blue, feature value is low. The y-axis (SHAP values) indicates in which direction that feature affects the prediction. That is, for control vs tumor classification task, a positive SHAP value indicates that feature for that sample was important to label it as a tumor sample. On the other hand, a negative SHAP value indicates the feature was important to label it as a control sample. For instance, many purple dots with high SHAP values indicate positive correlation between the tumor and the magnitude of the peak at that ppm. For benign vs malignant classification task, a positive SHAP value indicates malignant label and a negative SHAP value indicates benign label.

We show our results in Figure 3. Here, we only annotate the peaks in the SHAP values (most important in either direction) that reach an absolute SHAP value of 0.005. We use the metabolite database provided by Ruhland *et al*. for annotation of peaks. We only list the names of the metabolites which have a group that exactly match with the base of the peak region (i.e., is a subset of the peak region). Note that there are usually many metabolite groups overlap with such regions. To limit the number of candidate metabolites, we use such a stringent criterion. We also annotate the peaks of two well known cancer biomarkers 2-hydroxyglutarate and creatine.

**Fig. 3:**
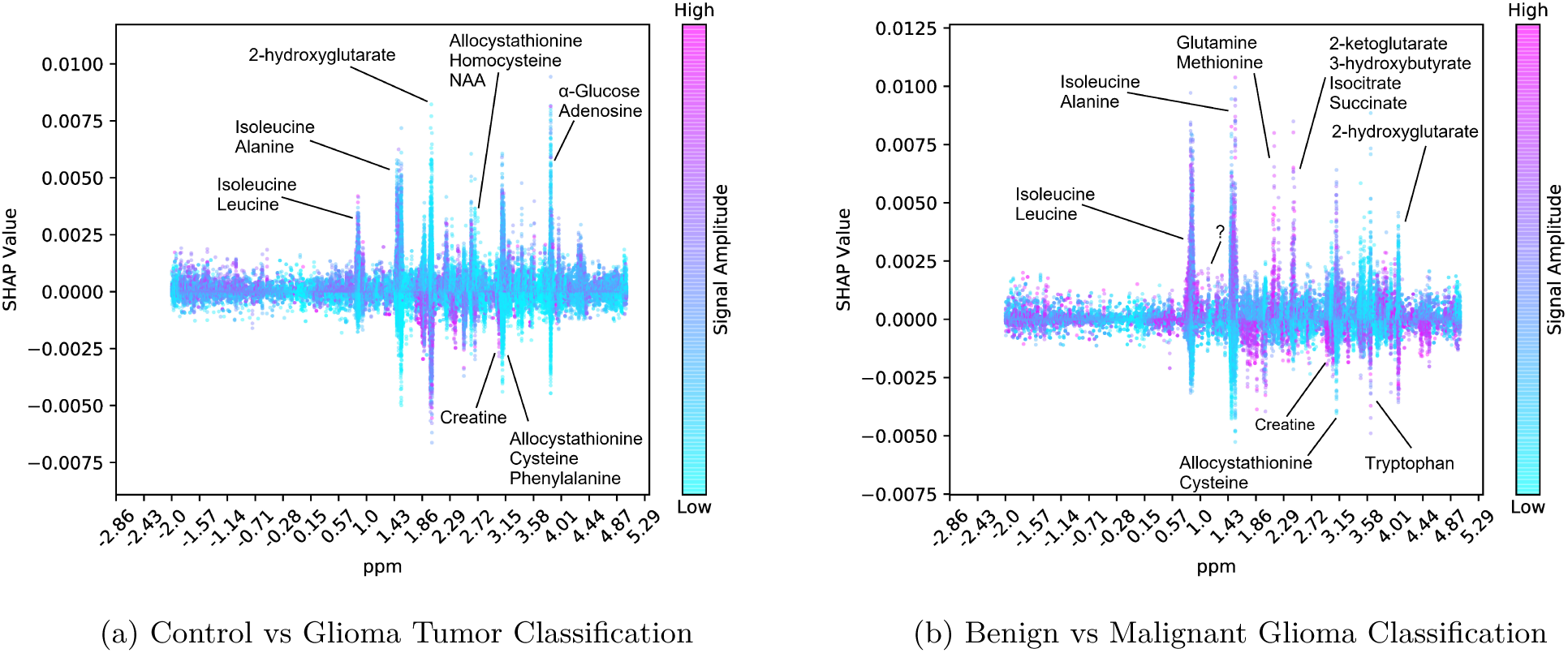
The SHAP Values (y-axis) for each ppm in the spectrum (x-axis) is shown for each sample (dots). Dot color purple indicates a high feature value, and blue indicates a low value. A positive SHAP value indicates that feature was important to classify that sample as (i) tumor as opposed to control in Panel A; and as (ii) malignant as opposed to benign in Panel B. Conversely, A negative SHAP value indicates that feature was important to classify that sample as (i) control as opposed to tumor in Panel A; and as (ii) benign as opposed to malignant in Panel B.

First, we find that 2-hydroxyglutarate has high feature importances in both classification tasks. Isocitrate Dehydrogenase (IDH) is a rate limiting enzyme in the Krebs cycle and plays an important role in the regulation of the energy metabolism. IDH mutations are known to affect tumor metabolism. For instance, Mutations of IDH are known to produce high levels of 2-hydroxyglutarate that inhibits glioma stem cell differentiation [12, 39]. So, low levels of 2-hydroxyglutarate indicate malignancy. In line with this information, we observe that when the corresponding peak (feature) values are low (i.e., blue), SHAP values are high which indicates that those samples are predicted to contain tumor and malignant cells, respectively. Similarly, creatine is a well-known biomarker for gliomas. Low creatine levels are observed in IDH mutants gliomas indicating low grade (benign) tumors [42, 43]. In both tasks, we observe blue peaks with high SHAP values for the ppm range that coincides with creatine groups. This indicates that when creatine levels are low, we predict the sample to be tumor and malignant, respectively. Thus, our model had learnt to focus on regions in the spectrum which are used by technicians today as indicators.

For both tasks, we consistently find that peaks belong to branched chain amino-acids isoleucine and leucine are focused by the model. These amino acids are known to have altered concentrations in the presence of IDH mutations, but their status as a biomarker for gliomas are not strongly established. We also observe that various other amino acids are also focused by the model as annotated in Figure 3. This suggests possible biomarkers due to the altered amino acid metabolism. Finally, while distinguishing benign and malignant gliomas, we observe that 2-ketoglutarate and Isocitrate are also important factors for successful classification. This is also meaningful as the IDH enzyme catalyzes the reaction that converts one to other in reversible fashion. IDH mutations affect this process and produce more 2-hydroxyglutarate from 2-ketoglutarate rather then to produce isocitrate [12]. Thus, these are also candidate biomarkers stressed in the prediction of the algorithm.

The interpretation of the results is limited by the 76 metabolites and their ppm signatures provided in [30]. We have performed an analysis to find any SHAP value peaks that are not associated with any metabolite. We obtained top 200 peaks out of 8, 172 and found a relatively short attention peak near 1.00 ppm which indicates malignancy when the concentration is high. This is an uncharacterized region and might suggest a new biomarker. Furhter research and validation is needed to establish an understanding of the compounds in those regions and their relation to tumor metabolism. Yet, this shows the potential for the untargeted analysis we propose here, as such regions are discarded by an human analyst.

## 4 Discussion

Using a machine learning approach in this application has advantages over a technician commenting on the presence or absence of known biomarker metabolites using the raw signal. Our current catalogue of metabolites in the 1H HRMAS NMR spectrum is limited which means we potentially discard valuable information with this targeted analysis. On the other hand, the RF algorithm we use generates decision tree classifiers, each of which focus on different parts of the spectrum and process features in combinations. Thus, the algorithm performs an untargeted analysis as there is no metabolite identification/quantification. The analysis is also non-linear and multivariate unlike the current approach based on one by one quantification of certain metabolites. Moreover, fluctuations in chemical shift is common in NMR results and a binary guess is needed to conclude whether a peak belongs to a certain metabolite. The RF model can average out such inconsistencies. As seen in Figure 3, the focus (i.e., given importance) of the algorithm resembles a peak around certain ppm regions, indicating a smooth adjustment of the weights associated with each ppm, according to the composition in the training cohort.

Our results provided in Section 3.2 show that our models achieve high AUC and AUPR values indicating that the RF is a viable method to be used in the surgery room. The average test time of the model is negligable (i.e., 0.01 secs.) which makes it possible to use it in real-time. The training phase is performed offline and on average takes 25.2 mins. We interpret the results of the RF model using SHAP values provided for each ppm in the spectrum. We validate that groups of known cancer biomarkers such as creatine and 2-hydroxyglutarate had an important role in the decision made by the model. This is an important feature for this analysis as usually a surgeon would like to know the reasoning behind the decision made by a program. We also indicate several ppm regions which have been important for the classifications. These regions harbor shared groups of several metabolites and further research is needed to validate their ties to glioma metabolism and their status as a glioma biomarker.

We observe that formulating the problem as a multi-class classification problem and trying to distinguish benign, malignant and control samples does not perform well. The number of benign samples is small and it is hard to distinguish them as their signal resembles the controls. Thus, the mean class AUCs we obtained for control and benign samples were down to 60% and 40%, respectively. malignant samples are successfully classified (mean AUC =90%) Since, the primary goal is to distinguish tumor and healthy tissue we opted for the presented scheme in this study.

While benchmarking several machine learning algorithms, we observe for both tasks that convolution operation improves the performance of the baseline neural network model slightly and has somewhat lower variance in the performance. Despite being edged by the RF model, we think CNN model can perform well when trained on larger datasets. Our dataset is, to the best of our knowledge, the largest cohort with close to 600 labeled samples. However, CNN uses a deep architecture and requires larger cohorts to learn more complex features. We would like to note that we performed extensive testing on the CNN architecture, which varied the number of layers, number of kernels, activation functions, pooling operations etc. We also experimented with a self-attention mechanism to find regions of interest in the spectrum. The results presented is the best set of results obtained for CNN model. We concluded that the model is too complex to be learnt with this sample size.

## 5 Conclusion

In this study, we developed a random forest based machine learning approach to distinguish glioma samples (benign or malignant) from the control samples using the 1H HRMAS NMR signal as the sole input. In our experiments, we show that the approach is efficient, accurate and interpretable. It can work in real-time and thus, can be used as a means of providing feedback to the surgeons on the left-over tumor samples during surgery.

## Data Availability

The data will be made available upon publication.

## Acknowledgements

This work was supported in part by grants from BPI France (ExtempoRMN Project), Hôpitaux Universitaires de Strasbourg, Bruker BioSpin, Université de Strasbourg and the Centre National de la Recherche Scientifique (CNRS). It is also supported by TUBA GEBIP award to AEC. We would like to acknowledge the helpful discussions of Furkan Ozden.

## Notes

### Competing Interest Statement

The authors have declared no competing interest.

